# Strengthening health service delivery and governance through institutionalizing ‘Urban Health Atlas’ - a geo-referenced Information Communication and Technology tool: lessons learned from an implementation research in three cities in Bangladesh

**DOI:** 10.1101/2022.03.24.22272880

**Authors:** Sohana Shafique, Dipika Shankar Bhattacharyya, Tarek Hossain, Shaikh Mehdi Hasan, Shakil Ahmed, Rubana Islam, Alayne M. Adams

## Abstract

**Introduction:** Urban health governance in Bangladesh is complex as multiple actors are involved and no comprehensive data are currently available on infrastructure, services, or performance either in public and private sectors of the healthcare system. The Urban Health Atlas (UHA) – a novel and interactive geo-referenced, web-based visualization tool was developed in Bangladesh to provide geospatial and service information to decision makers involved in urban health service planning and governance. Our objective was to study the opportunities for institutionalization of the UHA into government health systems responsible for urban healthcare delivery and document the facilitators and barriers to its uptake.

**Methods:** This mixed-methods implementation research was carried out during 2017-2019 in three cities in Bangladesh: Dhaka, Dinajpur and Jashore. During the intervention period, six hands-on trainings on UHA were provided to 67 urban health managers across three study sites. Both in-depth and key informant interviews were conducted to understand user experience and document stakeholder perceptions of institutionalizing UHA.

**Results:** Capacity building on UHA enhanced understanding of health managers around its utility for service delivery planning, decision making and oversight. Uptake of UHA was challenged by inadequate ICT infrastructure, shortage of human resources and general lack of ICT skill among the managers. Motivating key decision makers and stakeholders about the potential of UHA and engaging them from the inception helped the institutionalization process.

**Conclusion:** While uptake of UHA by government health managers appears possible with dedicated capacity building initiatives, its use and regular update are challenged by multiple factors at the implementation level. A clear understanding of context, actors and system readiness is foundational in determining whether the institutionalization of health ICTs is timely, realistic or relevant.

## Introduction

By the year 2040, half of the population of Bangladesh will reside in cities with almost one-third of urban dwellers living in informal settlements [1]. Although the country has made remarkable progress in reducing maternal and child mortality and achieving global goals [2], there are significant disparities in maternal neonatal and child health (MNCH)-related indicators and access to services across socioeconomic strata especially in urban areas [3–5]. Of particular note are higher rates of childhood stunting and infant mortality in urban informal settlements compared to other urban areas and the national average [5–7]. Persistent inequities in key health indicators stem from social determinants of health as well as deficits in health service coverage, access, and quality, including timely and appropriate primary to tertiary-level referral. These inequities have widened as a result of unprecedented COVID-19 related disruptions to health systems and to livelihoods of the urban poor [8–9].

The urban health system in Bangladesh is pluralistic and characterized by inadequate coordination and regulation, and geographic and socioeconomic inequities in healthcare access [10–13]. Poor planning and management capacity, lack of clear roles and responsibilities among various health-related authorities, and gaps in service coverage and key human resources are challenges in ensuring urban health [14]. The formal private sector is massive, accounting for 80% of over 3500 hospitals in Bangladesh, and an even greater percentage in urban areas [14]. Lack of regulation of this sector has resulted in concerns about the quality of care and financial accessibility, especially for the urban poor [15–16]. Inadequate public primary care infrastructure and services disproportionately impact the urban poor and pose a significant challenges to the country’s aspirations to meet the goal of Universal Health Coverage by 2030 [17–18].

The World Health Organization (WHO) suggests that all member countries have a master facility list to support health service delivery and monitoring [19]. Enabling the use of data for strategic decision-making and better governance, institutionalized health management information systems (HMIS) represent the foundation upon which improvements in health outcomes can be monitored and greater accountability ensured [20]. In this regard, the Ministry of Health and Family Welfare (MOHFW) in Bangladesh is implementing the District Health Information Software-2 (DHIS2) with support from development partners to facilitate evidence-informed planning and decision making [21]. While the system has been rolled out nationally, information is largely confined to public healthcare facilities. In urban areas, except for large public hospitals and some NGOs involved in primary care provision, data are particularly sparse, especially for the large private sector.

To address this information gap, a geo-referenced health facility database for nine major cities and municipalities across Bangladesh [22] was assembled. To make the data more accessible and useful to non-technical policy makers and public health managers [23], an Information Communication and Technology (ICT) tool called Urban Health Atlas (UHA) was developed (http://urbanhealthatlas.com). This Geographic Information System (GIS) based interactive online tool displays health facility data visually and permits their manipulation for better healthcare planning and decision making. Providing detailed information on the location and services available at public and private health facilities, it allows users to examine gaps and duplication in service provision, assess the coverage of emergency services and the availability of doctors in 24 hours, visualize population coverage and poverty mapping, calculate the shortest distance to referral facilities from any location, and determine whether a given facility is licensed and registered.

In the present context of urban health in Bangladesh, UHA could serve as an important tool to support health planning processes. The study aims to document stakeholder perceptions and experiences in adopting this tool to assist in strategic healthcare planning, day-to-day decision-making and oversight, with the goal of increasing equity and efficiency in urban MNCH services.

## Methods

### Study design and participants

This implementation research [IR] was conducted between 2016 and 2019 and employs a mixed-methods approach [24]. Detailed methods have been reported elsewhere [25]. Briefly, the WHO PATH toolkit was used to guide the introduction and implementation of information and communications technology (ICT) in health information systems [26].

### Study sites

Two city corporations (CCs) – Dhaka North City Corporation (DNCC) and Dhaka South City Corporation (DSCC) – and two municipalities, Jashore and Dinajpur in northern and south divisions were purposively selected, both of which have geo-referenced health facility data collected by icddr,b.

### Sample size

Through a stakeholder mapping exercise, national and local level urban health decision makers were engaged to help identify potential users for UHA, yielding a sample of 15-16 respondents for each of 4 study sites. The sampling strategy, type, and number of respondents in each site are published elsewhere [25].

### Implementation process

The study implementation process began with stakeholder sensitization and partnership-building through two consultation workshops. These workshops served to generate suggestions on how to promote uptake, regular use, and update of UHA, create buy-in for the implementation research project, and promote stakeholder engagement in UHA advocacy. As a result of this process, an implementation partnership with the Management Information System (MIS) of the Directorate General of Health Services (DGHS), MOHFW was formalized through a Memorandum of Understanding (MoU) between icddr,b and DGHS. Permission letters were also obtained from mayors of CCs and municipalities.

The UHA training intervention consisted of capacity building sessions for government and other urban health planners and managers on how to employ UHA as a planning tool to integrate and expand maternal, newborn, and reproductive health services with a focus on improving access for the urban poor. Trainees were selected in consultation with local government authorities. The total number of participants enrolled in capacity-building training by city is shared in Table 1. All institutional agreements and permissions were sought in advance from local government institutions and the health ministry.

**Table 1.**
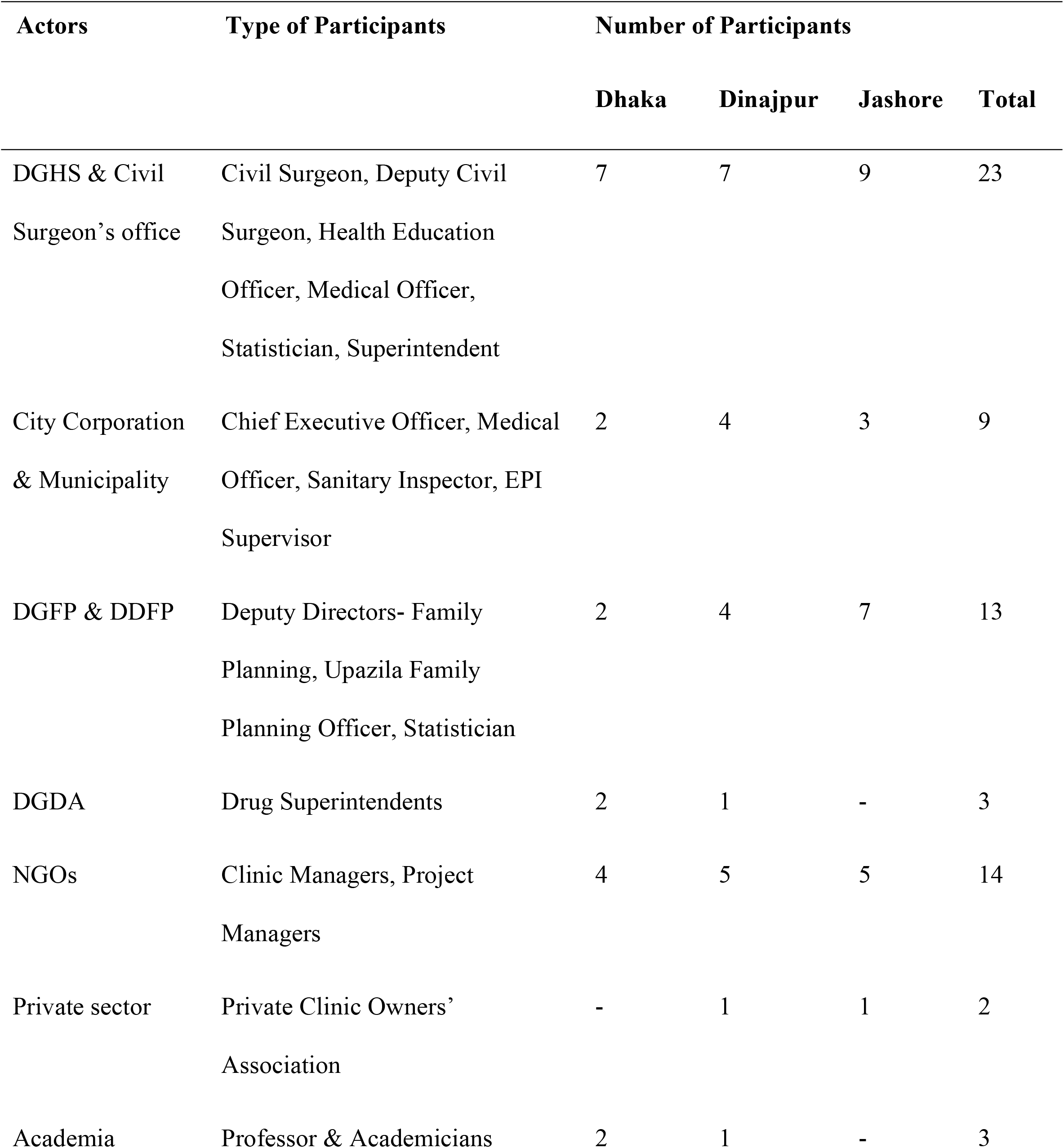
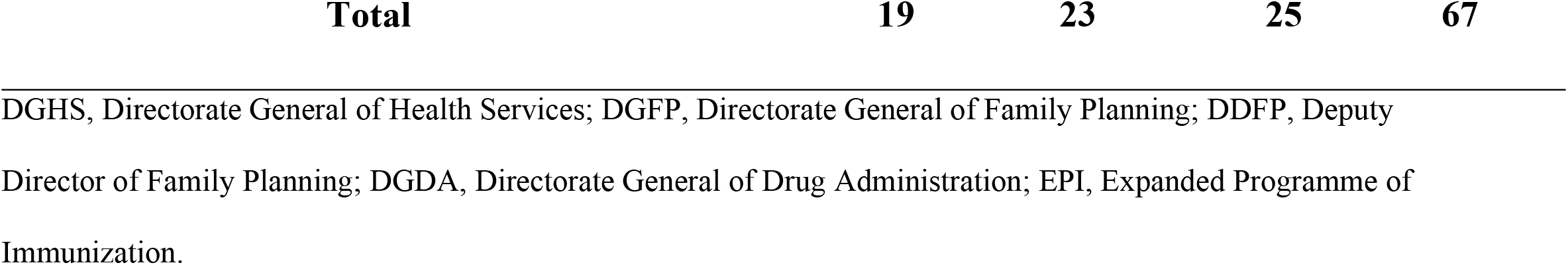
Total number of participants in the capacity building workshops in Dhaka, Dinajpur and Jashore.

Capacity building was divided into two phases: In the first phase, a two-day training was provided to participants that involved a series of hands-on exercises; and in the second phase, a refresher training was conducted three months post basic training among the same participants. A training manual was prepared to guide UHA capacity-building workshops including case studies, guidelines for group work and hands-on activities, pre-test/post-test questionnaires, etc. The training was organized with key actors in the urban healthcare system including health workers and managers working in local government, and NGOs. Training modalities were contextualized based on local needs in discussion with implementation partners, and facilitated by experts in urban health, and health management and geographic information systems (HMIS and GIS). Training sessions provided an introduction to current urban health challenges in each city, followed by an overview and demonstration of the UHA and its functions. Hands-on training, group work, and case studies were undertaken to familiarize users with UHA and to get their feedback on how it could be improved to better meet their needs, and suggestions on how health services information could be updated.

### Data collection

Methods used for data collection included desk review and a series of in-depth interviews conducted by an experienced group of researchers with expertise in qualitative and mixed-methods approaches. The desk review involved the critical assessment of existing policies related to urban health and the use of ICTs in for primary care and MNCH decision making, including the: i) 2011 National Health Policy; ii) The 2009 National ICT policy; iii) 2014 National Urban Health Strategy; iv) Program Implementation Plan of MOHFW. Interviews were conducted with stakeholders and trainees prior to, during and post the UHA intervention employing the steps for ICT institutionalization outlined in the WHO PATH toolkit. Key informant interviews (KIIs) with urban health system actors and policymakers along with desk reviews were undertaken to document how MNCH planning and referral decisions are currently made. In-depth interviews (IDIs) with potential UHA users were conducted to explore user preferences and information needs to refine the UHA tool in advance of the training. A subsequent set of IDIs with UHA users sought to understand their experiences and to document challenges and successes of using UHA for MNCH service decision-making during training and one- & three-months post-training. Alongside interviews, process documentation of the dynamics of UHA uptake was undertaken, which considered contextual factors and their influence. All interviews were recorded with participant consent, but with simultaneous note-taking in case of equipment failure.

### Data analysis

Data transcription occurred immediately following each interview, followed by translation into English. Data familiarization involved reading the transcripts repeatedly to appreciate data content, structure, and emerging themes. Initial analyses involved developing a process flowchart for current decision-making practices using the KII results and organizational process reviews. In addition, a list of user needs was extracted that helped identify the most important and feasible functions to be added to UHA. Analysis of UHA user interviews involved applying “a priori” codes derived from the WHO PATH toolkit [WHO/PATH], and exploring patterns and themes using the Framework Method [27].

Transcripts were coded using ATLAS-ti (version-7.5.7). A team approach to analysis was employed to minimize individual biases and periodic inter-coder reliability checks were organized. Group discussions of emerging themes and patterns in the data were oriented around data displays that allowed for more systematic pattern-testing and cross-checking across respondents.

### Ethics

The study protocol was approved the Research Review Committee (RRC) and the Ethical Review Committee (ERC) of icddr,b, both of which provided a thorough and critical review of its technical and ethical aspects. Participants were asked for written consent before interviewing and were kept anonymous and unidentifiable. All data were kept in locked storage, or controlled access folders, allowing only investigators of the study and members of the ERC of icddr,b to access information, if needed.

## Results

Three key themes related to the ‘Uptake’, ‘Use’ and ‘Update’ of the UHA were identified. Within the area of ICT use, the concepts of *understandability, usability*, and *utility* were also explored. *Understandability of UHA* refers to the extent to which the tool’s features are understood by users *Usability* denotes the extent to which the tool and its features are easy to use and apply, while *utility* captures the tool’s perceived practical application by users. Each of these themes are discussed here, with a focus on factors facilitating or challenging the institutionalizing process of UHA at both individual and organizational levels (**Fig 1**).

**Fig 1.**
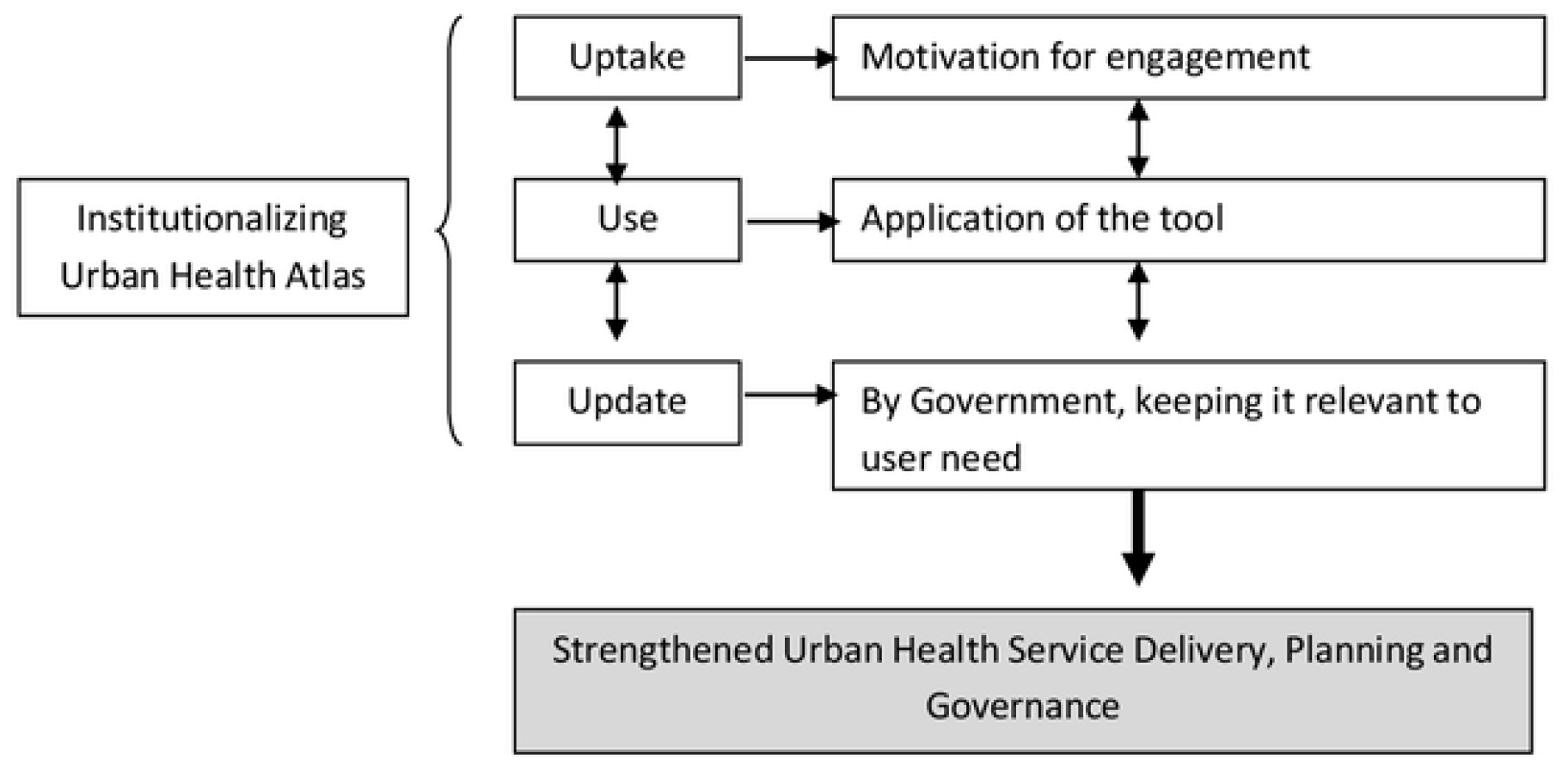
Original conceptual framework for institutionalization developed by authors before implementation.

### Factors influencing *‘uptake’* of UHA

As regards uptake of the tool, our findings indicated the importance of policy level support, and stakeholder involvement as crucial facilitators along with efforts to understand and respond to user needs and responsibilities. A major challenge was the degree of individual, organizational and system readiness for a digital tool and its application in the absence of formal systems of urban health service planning.

#### Policy-level support for ICT and evidence-informed planning and decision making

Bangladesh has made remarkable progress in terms of incorporating ICT-based innovation into the health system with policy support [21]. The country is now one of the largest implementers of the Demographic Health System 2 (DHIS2) globally, with an average reporting rate of 98% [28]. A number of other digital platforms and software for collecting, analyzing, and getting feedback on routine health-related data are also in place [29]. From the policy review, it was found that the Health Ministry’s (MOHFW) commitment to improving health outcomes and producing and digitizing real-time health data are factors supportive of ICT innovation and uptake in rural health systems including tertiary-level hospitals. However, within urban areas where responsibility for primary care services falls with the Ministry of Local Government, Rural Development and Co-operatives (MOLGRD&C), investments in digitized platforms and processes, including regular capacity strengthening programmes for managers around routine HMIS, have been less apparent. Thus, while policy appetite for ICT exists at the national level, this has not been translated into ICT uptake at the local government level especially in the context of urban service provision.

#### Involving stakeholders from the beginning

Rapport building with key stakeholders in the early stages of the project and the development of formalized institutional agreements, were fundamental to the implementation process. A letter of support was issued by the Director, MIS, DGHS under the MOHFW ensuring their commitment to facilitate project implementation as appropriate, while MoU’s and permission letters from local level authorities helped mitigate bureaucratic challenges. These efforts helped ensure the cooperation of health workers and managers within local government institutions and the DGHS, and facilitated project roll-out from design to implementation.

To further cultivate stakeholder interest in shaping the implementation of UHA within the government system, a Technical Advisory Group (TAG) was formed consisting of urban health actors including Government, development partners, NGOs, academicians and public health and e-Health experts. The Director General, DGHS assumed the role of Chair. TAG members were purposively selected with the goal of creating well-placed advocates for UHA and its uptake for urban health decision making, planning, referral and regulation at the policy level. Three TAG meetings were organized throughout the study period. In addition, two stakeholder consultation workshops were arranged to explore their interests, roles, information needs and perceptions on how UHA might benefit their respective institutions. Useful suggestions were generated on potential institutionalization processes for UHA, and how to make the UHA user-friendly for health managers. For example, the filter searching options in UHA were simplified and a Bangla interface was added based on suggestions received by senior government officials.

#### Assessing system readiness for institutionalizing UHA or any ICT tool

System readiness greatly influenced uptake of UHA like any other ICT tool. At the institutional level, lack of available infrastructure and training, and bureaucratic barriers to obtaining both, were the major challenges regarding the uptake of UHA. Infrastructural capacity constraints were widespread within local government and the district office of the Ministry of Health, including inadequate internet supply, ICT equipment and human resources. Local-level managers also reported bureaucratic delays in procuring and repairing equipment, and renewing software licensing, as well as a lack of needs-based ICT training.

Beyond institutional level challenges, individual barriers to adopting ICT tools were noted. For example, one informant from the DGHS, MOHFW commented that even after issuing government orders for ICT uptake from the central management level, it took a long time for people to become comfortable with new ICT tools. There was also resistance against replacing familiar systems with unfamiliar technology. In his opinion:

> *“In any organization, resistance (to adopt a new ICT tool) is high… many senior staff are reluctant to use it as they have been using the manual system for years and years*.
>
> *Therefore, reluctance to change in the system might be a barrier (for institutionalizing UHA)*.*” (KII 1, DGHS, Dhaka)*

#### Tailoring UHA to enable the regular assigned tasks of potential users

We documented the tasks and processes managers need to accomplish their job responsibilities and how UHA might assist in performing these tasks more efficiently. One NGO programme manager mentioned that knowing about the distribution of facilities and their service details across the urban areas was helpful carrying out their duties, while a government official noted:

> *“We do not have well-organized data on private sector facilities. As a result, we missed some information, especially in terms of institutional delivery and MNCH services provided by private sector… if we can make a unified system to report who provided the service and how they provided this, it will be helpful in executing our job. The UHA can help the coordination process*.*” (IDI 3, government official, Jashore)*

#### Prior experience with data-driven planning and decision making

Implementation of various ICT based tools in the health sector (e.g. DHIS2, HRIS, OpenMRS, Tableau etc.) have facilitated data-driven decision making. It is not surprising, therefore, that prior experience with government ICT systems was beneficial in understanding the potential of UHA and influenced levels of comfort with the tool. It should be noted, however, that exposure to and comfort with ICTs varied substantially by age, and that internet literacy also varied by cadre among government officials and managers.

#### Perceived utility by potential users

The most commonly perceived application of UHA was in enabling planning-related tasks and performance given its comprehensive geo-coded database of health facilities. Apart from this organizational advantage, study participants noted individual benefits. These related to its potential personal use in helping search for a specialist or the nearest facility during an emergency situation. Many suggested that the existing web-based version of the UHA tool might be adapted as a mobile application to enable offline access to data and greater convenience.

#### Capacity building on UHA & modification of the tool

Hands-on capacity building workshops and refresher trainings were appreciated by participants and served to identify and resolve practical issues affecting the usability and understandability of UHA. One such issue was the slow loading time of the tool, which was subsequently reduced by by separating out drug shops from health facilities (e.g. hospital, clinic, NGOs etc.) in the search function. The continuous refinement of the tool and tailored modifications in response to user needs and demands are important facilitating factors for institutionalizing any ICT.

### Factors influencing *‘use’* of UHA

We investigated the use of UHA under three major sub-themes: *Understandability, Usability*, and *Utility*. Factors facilitating or challenging the understandability of UHA among users in the study were training, compatibility with work, and previous exposure to ICT tools or e-health technologies. Participation in organized hands-on training was the most cited reason for improved understanding of the tool. For users who had previous exposure to ICT and had worked with similar ICT tools, the UHA was easily understood and its design and features were well-recognized. The categorization of data in UHA by type of facility, defined area location, price list and opening hours helped to make the UHA interesting to participants and facilitated understandability of its various functions.

Efforts to increase usability through a Bangla translated version of the tool, new filter options, and training around these features were especially appreciated. As one NGO manager said:

> *“Through the training on UHA, we got to know where we need to click to find the kind of data we need and made it useful for us…the whole urban health scene of Dinajpur (is) in one place - hat my organization is doing, what the government is doing, what other health facilities are doing…” (IDI 5, NGO Personnel, Dinajpur)*

The utility of the tool was mainly perceived in the area of planning, decision making and oversight by managers, especially in terms of redistributing MNCH facilities more equitably by geography and population needs. According to the district health manager of one municipality:

> *“The information provided in UHA: the location, the ward boundaries, and the corresponding number of facilities will help urban health planning purposes. For example, now I can see that certain wards contain many facilities, whereas other wards have few. So I can redistribute them, and plan and implement in a more (evidence-based) way. If implementation occurs, I can use these maps to distribute and supervise my workers and decide where I need more support. It will also help me in evaluation to understand what things were supposed to be done what needs to be improved*.*” (IDI 1, Government Official, Dinajpur)*

Study participants opined that one of the strengths of UHA is its complete database of healthcare facilities, including their geo-location and related service details. As such, participants thought it was a unique resource with many possible applications with respect to MNCH planning in complex urban settings. In fact, these applications were used by health planners to improve the distribution of MNCH health services in Dinajpur [30] by identifying and reducing duplication and service gaps among satellite centers and Extended Program on Immunnization (EPI) sites and ensuring that designated service providers were accountable for delivering services in designated wards. The tool was also used for service re-allocation and emergency health service management in Dinajpur [30]. Using ward-wise population data available in UHA, decisions were made regarding emergency needs for Oral Rehydration Solution (ORS), zinc tablets and other drugs:

> *“During the last flood in Dinajpur, I estimated the ward-wise population from this tool. This helped me to estimate how many zinc tablets and other emergency drugs needed to be distributed (per ward). When I will present the data of my city during this year’s flood, I can demonstrate it using UHA*.*” (IDI-6, Dinajpur)*

As patients and service seekers themselves, study participants also recognized the tool’s worth from the demand side, and its potential in helping access appropriate and proximate care while minimizing the involvement of middlemen.

### Factors facilitating regular *‘update’* of UHA

Lack of an agreed mechanism to update and link urban facility data to routine HMIS data, was perceived as an impediment to regular use of UHA for planning purposes. Weak administrative and planning capacity at local level and a limited urban mandate at the national level accounted for this absence. As one study participant explained:

> *“It is a useful tool but there is a lot of scope for improvement. The most important one is regular updating of facility and service data. There should be proper mechanisms and guidelines on how, when, and who should update it*.*” (IDI 3, government official, Jashore)*

High turnover of those in key decision-making positions was perceived to further hinder or delay UHA update or institutionalization. Shifting priorities, lack of interest and commitment, and differences of opinion on how and who should take charge of update, were widespread. For example, in Dinajpur, stakeholders stated that the Civil Surgeon’s office should be responsible for updating UHA information since all health-related reports are the responsibility of the Civil Surgeon’s Office. On the other hand, in Jashore, stakeholders opined that municipal authorities should lead the process of regular update given that urban primary health care falls under the jurisdiction of urban local body of MOLGRD&C. In both cases, however, there was a common understanding that a separate unit was needed to ensure accountability and coordination among those collecting, providing and using the information available in UHA. Regular capacity building would also be needed.

The conceptual framework was revised considering the findings of the study (Fig 2).

**Fig 2.**
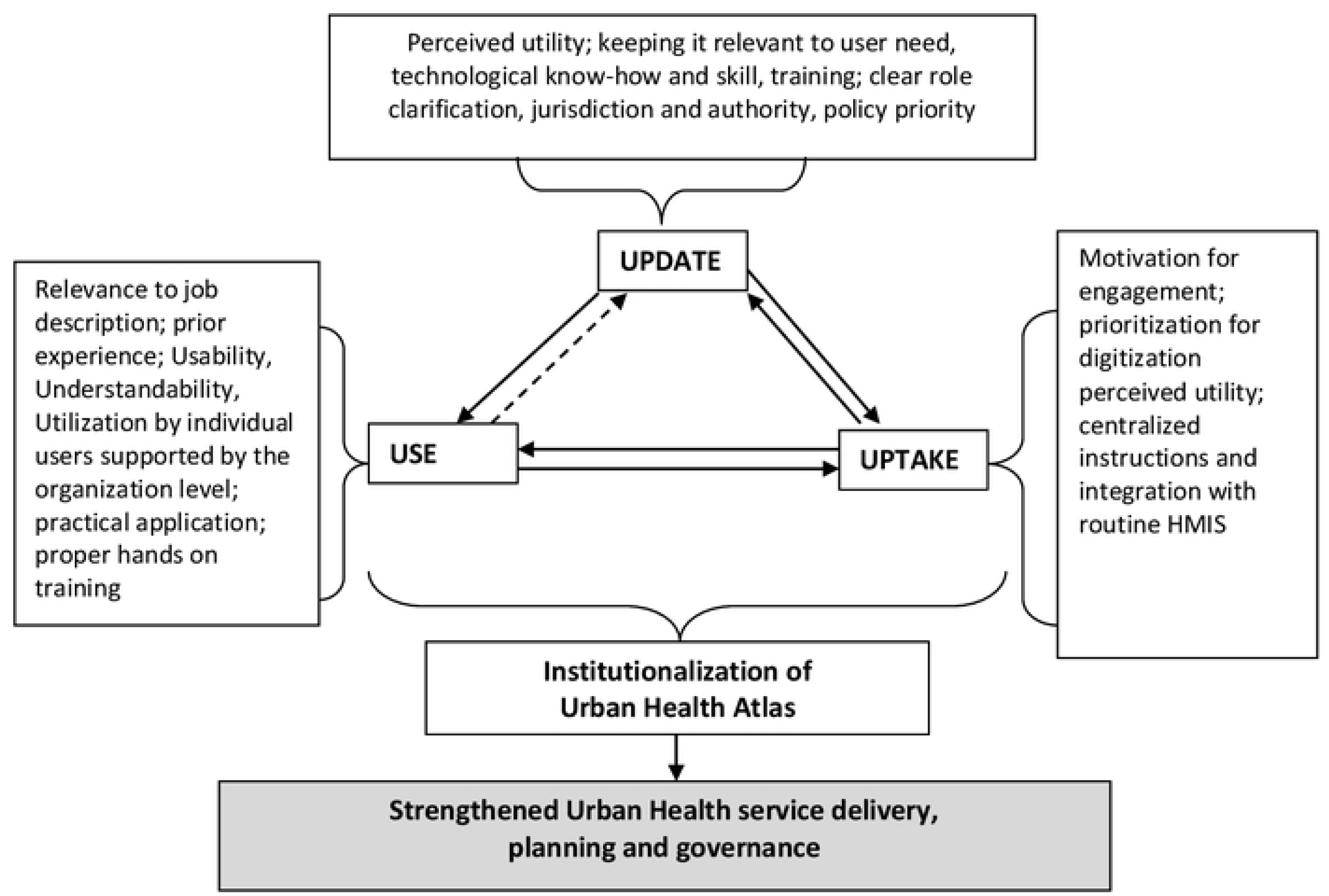
Revised conceptual framework for institutionalization of Urban Health Atlas.

## Discussion

This mixed-method implementation research is among the first studies in Bangladesh to explore processes for institutionalizing geo-referenced ICTs to strengthen health service planning, referral, and oversight in urban areas. Study findings demonstrate the utility of geo-referenced health facility information for MNCH service planning and decision making in urban settings and the value of data visualization as a means of enabling data use by non-technical persons. Findings also emphasize the necessity of capacity strengthening around ICT use to support evidence-based decision making. Despite the potential of UHA for urban health planning, many factors hindering ICT uptake, use and update were identified which undermine the goal of institutionalization. At the same time, the value of integrating and scaling up urban health facility information into the national health systems was recognized, as well as key implementation strategies.

Study results reinforced the importance of engaging stakeholders at the project conceptualization phase to facilitate implementation and ensure that ICT products are relevant and feasible. Bryson et al [31] similarly emphasized early stakeholder engagement as crucial to the development of ICT tools that are technically feasible and politically acceptable, while Makinde *et al*., [32] attributed ICT adoption to the involvement of multilevel stakeholders, from users to policymakers. In the context of Bangladesh, an engaged, multi-stakeholder approach was particularly crucial given the complexity of urban health service delivery involving multiple state and non-state authorities and actors at implementation and policy-making levels [33]. Indeed, given the pluralistic nature of many urban health systems in LMICs, multi-sector, multi-level engagement around ICT uptake and service delivery planning is essential in efforts to increase equitable access to healthcare [18, 34].

Consistent with our results, Mengiste et al. [35] identified lack of infrastructure as a key factor limiting system readiness for the adoption of ICT tools in the health sector in LMIC settings. This is supported by a study of DHIS2 deployment in Bangladesh, which noted poor internet connectivity and irregular electricity as hindering implementation [36]. These issues of infrastructural barrier are accompanied by resistance to change at individual and/or organizational level which is typical of many government bureaucracies in LMICs [37].

In terms of individual-level factors influencing uptake of ICT, our study identified the perceived utility of the tool, and the user’s job description, and professional capacity as especially important. This finding is congruent with a systematic review conducted by Gagnon et al. [38] that highlighted perceived ICT usefulness, familiarity and appropriate training of end users as the main factors influencing adoption. To this point, our results indicated the importance of understanding job-related needs and preferences of potential users prior to ICT implementation. Modifications to the tool such as introducing the Bangla interface and adapting filtering options to better reflect user needs, improved its acceptability and facilitated both policy buy-in and uptake and use of UHA. These observations are consistent with a literature suggesting that adoption of eHealth innovations is unlikely if the existing skills and capacities of target users are insufficient [38–39]. One scoping review examining factors enabling ICT uptake and scale, emphasized the importance of technical training, demonstrating the value of the innovation, and customizing the technology in terms of local language and user culture and comfort, including interface appearance [40]. Likewise, in Tanzania, it was found that the introduction of a tool intended to improve the delivery of health care services was only possible when there was a perceived need for the tool and prerequisite ICT training [41].

When examining the adoption or ‘use’ of our ICT tool we considered three dimensions - Understandability, Usability, and Utility – each of which influenced the extent to which it might be employed by users and the institutions in which they work. Successful use of ICT requires understanding, or more specifically digital literacy. In South Asia, the lack of well-trained ICT personnel was found as a barrier to the adoption of health technology [42]. Contrariwise, in Ghana, internet literacy improved confidence in using ICT tools in service delivery [43].

The usability of ICT is also critical. A systematic review in low-and middle-income countries (LMICs) identified that faulty, fragmented, and limited functionality of electronic health systems and software developed in languages other than English, were the main technical barriers to adopting ICT tools in LMICs [44]. In terms of utility, ICT systems that are not a good fit with work practices or regular activities are reported as barriers for successful implementation [38]. While health workers in Uganda perceived ICTs as beneficial for job performance; limited knowledge and skills, poor internet networks, and inadequate logistics supply were the reported barriers to use [45]. Likewise, participants in our study considered UHA as beneficial and relevant to their daily assigned tasks, yet, lack of infrastructure, logistic support and system readiness hampered routine implementation.

Despite these challenges, the use of ICT tools for health systems planning has grown exponentially [29, 46–47]. Participants in our study perceived many potential applications of UHC in health service planning, decision making, as well as facility and emergency management and its benefits in terms of efficiency and accountability. For example, the adoption of UHA in Dinajpur district as a tool for planning, assisted in decisions about the location of NGO services to improve service coverage.

Regardless of setting, a key requirement of institutionalizing ICT is maintenance of quality, up-to-date data. In implementing a health workforce planning tool in the USA, collaboration between state agencies, stakeholders and relationships among data contributors and data stewards were needed to keep the application current [46]. In the context of UHA, participants emphasized that institutionalization was contingent on keeping health services data up-to-date and identifying a responsible authority for this purpose.

### Limitations

The study took place in two city corporations and two municipalities in Bangladesh where UHA had been implemented. Given the extraordinary diversity of urban settings, generalizability may be limited. It is possible that in cities with different geo-political administration arrangements, such as federal and state level divisions in governance, different challenges may be faced.

Notwithstanding these differences, insights arising from this study enrich the literature on conditions, features and processes of successful ICT institutionalization.

## Conclusions

Institutionalization processes for health-related ICTs are influenced by multiple national, local and individual level factors which must be considered in planning for a successful and sustainable result. To enable uptake of UHA or any ICT into the government system, efforts to facilitate use of evidence in decision-making are prerequisite. These include developing necessary capacity and infrastructure and identifying or defining jobs that enable their useful application. A clear understanding of context, actors and system readiness is foundational in determining whether the institutionalization of health ICTs is timely, realistic or relevant. When institutionalized successfully, ICTs such as the UHA have the potential to strengthen government HMIS, enable governance and accountability and ultimately, help achieve Universal Health Coverage and sustainable development. In the context of this study, necessary next priorities for research is identifying feasible methods to keep the UHA up-to-date with the collaboration of the Ministry of Health and municipal services.

## Data Availability

All relevant data, underlying the results, are included in the manuscript. The detail qualitative transcripts can be found in icddr,b data repository. As per icddr,b data policy (http://www.icddrb.org/policies), access to these datasets is subject to reasonable request. Interested parties may contact Ms. Armana Ahmed, Head, Research Administration, icddr,b for inquiries related to data access.

## List of Abbreviations

ADB: Asian Development Bank
CCs: City Corporations
DDFP: Deputy Director of Family Planning
DGFP: Directorate General of Health Services
DGHS: Directorate General of Health Services
EPI: Expanded Program on Immunization
ERC: Ethical Review Committee
GIS: Geographic Information System
HMIS: Health Management Information System
ICT: Information Communication and Technology
IDI: In-depth Interviews
KII: Key Informant Interviews
LMICs: Low and Middle Income Countries
MNCH: Maternal, newborn and child health
MOHFW: Ministry of Health and Family Welfare
MOLGRDC: Ministry of Local Government, Rural Development and Cooperatives
NGOs: Non-Government Organizations
PHC: Primary Health Care
RRC: Research Review Committee
SDGs: Sustainable Development Goals
UHA: Urban Health Atlas
UHC: Universal Health Coverage
WHO: World Health Organization

